# Estimating the COVID-19 Infection Rate: Anatomy of an Inference Problem

**DOI:** 10.1101/2020.04.15.20066811

**Authors:** Charles F. Manski, Francesca Molinari

## Abstract

As a consequence of missing data on tests for infection and imperfect accuracy of tests, reported rates of cumulative population infection by the SARS CoV-2 virus are lower than actual rates of infection. Hence, reported rates of severe illness conditional on infection are higher than actual rates. Understanding the time path of the COVID-19 pandemic has been hampered by the absence of bounds on infection rates that are credible and informative. This paper explains the logical problem of bounding these rates and reports illustrative findings, using data from Illinois, New York, and Italy. We combine the data with assumptions on the infection rate in the untested population and on the accuracy of the tests that appear credible in the current context. We find that the infection rate might be substantially higher than reported. We also find that the infection fatality rate in Illinois, New York, and Italy is substantially lower than reported.

## 1. Introduction

It is well appreciated that accurate characterization of the time path of the coronavirus pandemic has been hampered by a serious problem of missing data. Confirmed cases have commonly been measured by rates of positive findings among persons who have been tested for infection. Infection data are missing for persons who have not been tested. It is also well-appreciated that the persons who have been tested differ considerably from those who have not been tested. Criteria used to determine who is eligible for testing typically require demonstration of symptoms associated with presence of infection or close contact with infected persons. This gives considerable reason to believe that some fraction of untested persons are asymptomatic or pre-symptomatic carriers of the COVID-19 disease. Presuming this is correct, the actual cumulative rate of infection has been higher than the reported rate.

It is perhaps less appreciated that available measurement of confirmed cases is imperfect because the prevalent nasal swab tests for infection are not fully accurate. It is important to distinguish nasal swab tests from serological tests. Nasal swab tests are used to detect the presence of live virus within a person, signaling an active infection. Serological tests are used to detect the presence of antibodies that the immune system develops after onset of infection. The presence of antibodies signals that a person was infected at some past date and shows that the body has subsequently produced detectable antibodies.

There is basis to think that accuracy of nasal swab tests is highly asymmetric. Various sources suggest that the positive predictive value (the probability that, conditional on testing positive, an individual is indeed infected) of the tests in use is close to one. However, it appears that the negative predictive rate (the probability that, conditional on testing negative, the individual is indeed not infected) may be substantially less than one. Presuming this asymmetry, the actual rate of infection has again been higher than the reported rate.

Combining the problems of missing data and imperfect test accuracy yields the conclusion that reported cumulative rates of infections are lower than actual rates. Reported rates of infection have been used as the denominator for computation of rates of severe disease conditional on infection, measured by rates of hospitalization, treatment in intensive care units (ICUs), and death. Presuming that the numerators in rates of severe illness conditional on infection have been measured accurately, reported rates of severe illness conditional on infection are higher than actual rates.

On March 3, 2020 the Director General of the World Health Organization (WHO) stated:^1^ “Globally, about 3.4% of reported COVID-19 cases have died.” It is tempting to interpret the 3.4% number as the actual case-fatality ratio (CFR). However, if deaths have been recorded accurately and if the actual rate of infection has been higher than the reported rate, the WHO statistic should be interpreted as an upper bound on the actual CFR on that date. Recognizing this, researchers have recommended random testing of populations as a potential future method to solve the missing data problem.^2^

In the absence of random testing, various researchers have put forward point estimates and forecasts for infection rates and rates of severe illness derived in various ways. Work performed by separate groups of epidemiologists at the Imperial College COVID-19 Response Team and the University of Washington’s Institute for Health Metrics and Evaluation has received considerable public attention.^3^ The available estimates and forecasts differ in the assumptions that they use to yield specific values. The assumptions vary substantially and so do the reported findings. To date, no particular assumption or resulting estimate has been thought sufficiently credible as to achieve consensus across researchers.

We think it misguided to report point estimates obtained under assumptions that are not well justified. We think it more informative to determine the range of infection rates and rates of severe illness implied by a credible spectrum of assumptions. In some disciplines, research of this type is called sensitivity analysis. A common practice has been to obtain point estimates under alternative strong assumptions. A problem with sensitivity analysis as usually practiced is that, in many applications, none of the strong assumptions entertained has a good claim to realism.

Rather than perform traditional sensitivity analysis, this paper brings to bear econometric research on partial identification. Study of partial identification analysis removes the focus on point estimation obtained under strong assumptions. Instead it begins by posing relatively weak assumptions that should be highly credible in the applied context under consideration. Such weak assumptions generally imply set-valued estimates rather than point estimates. Strengthening the initial weak assumptions shrinks the size of the implied set estimate. The formal methodological problem is to determine the set estimate that logically results when available data are combined with specified assumptions. See Manski (1995, 2003, 2007) for monograph expositions at different technical levels. See Tamer (2010) and Molinari (2020) for review articles.

Considering estimation of the cumulative infection rate for the coronavirus, we combine available data with credible assumptions to bound the infection rate. We explain the logic of the identification problem and determine the identifying power of some credible assumptions. We then report illustrative set estimates.

We analyze data from Illinois, New York, and Italy, over the period March 16 to April 24, 2020. We impose weak monotonicity assumptions on the rate of infection in the untested sub-population to draw credible conclusions about the population infection rate. We find that the cumulative infection rate as of April 24, 2020, for Illinois, New York, and Italy are, respectively, bounded in the intervals [0.004, 0.525], [0.017, 0.618], and [0.006, 0.471]. Further analyzing the case-fatality ratio, we find that as of April 24 it can be bounded, for Illinois, New York, and Italy, respectively, in the intervals [0, 0.033], [0.001, 0.049], and [0.001, 0.077]. The upper bounds are substantially lower than the death rates among confirmed infected individuals, which were, respectively, 0.045, 0.059, and 0.134 on April 24.

The analysis of this paper can be adapted to bound statistics related to but distinct from the cumulative infection rate in a population. One such statistic that may be of public health interest is the daily rate of new infection. Another is the fraction of the population who are currently ill.

We focus on the cumulative infection rate, and henceforth refer to it without the qualifier “cumulative.” Knowledge of this statistic is essential to forecast the level of herd immunity that a population has achieved by a certain date. It is also necessary to calculate probabilities of severe illness conditional on infection, including risks of hospitalization, treatment in an intensive care unit, and death. Knowledge of these probabilities is vital to inform both personal risk assessment and public health policy.

## 2. Methods

We first address the basic problem of bounding the infection rate as of a specified date. The analysis initially considers the problem abstractly and then derives bounds under particular credible assumptions. We next show how bounding the infection rate yields a bound on the rate of severe illness conditional on infection. We then extend the analysis to bound the rates conditional on observed patient characteristics. Knowledge of all of these rates is important to inform public health policy.

### 2.1. Bounding the Population Infection Rate

Consider a specified population of persons who are alive at the onset of the pandemic. This may, for example, be the population of a city, state, or nation. Let the objective be to determine the fraction of the population who are infected by the SARS-CoV-2 virus by a specified date d. Synonymously, this is the fraction of the population who experience onset of the COVID-19 disease by date d.

The present analysis assumes that a person can have at most one disease episode. If a person has been infected, the person either achieves immunity after recovery or dies. The assumption that immunity is achieved after recovery, at least for some period of time, is consistent with current knowledge of the disease. We also assume that the size of the population is stable over the time period of interest. Thus, we abstract from the fact that, as time passes, deaths from the disease and other causes reduce the size of the population, births increase the size, and migration may reduce or increase the size on net.

Let C_d_ = 1 if a person has been infected by the coronavirus by date d and C_d_ = 0 otherwise. The objective is to determine P(C_d_ = 1), the probability that a member of the population has been infected by date d. Equivalently, P(C_d_ = 1) is the infection rate or the fraction of persons who have experienced disease onset. Our analysis of the problem of inference on P(C_d_ = 1) is a simple extension of ideas that have been used regularly in the literature on partial identification, beginning with study of inference with missing outcome data (Manski, 1989).

P(C_d_ = 1) is not directly observable. However, population surveillance systems provide daily data on two quantities related to P(C_d_ = 1). These are the rate of testing for infection and the rate of positive results among those tested. To simplify analysis, we assume that a person is tested at most once by date d. This assumption may not be completely accurate, for reasons that will be explained later. In Section 2.2, we discuss the implications of its possible failure for our analysis.

Let T_d_ = 1 if a person has been tested by date d and T_d_ = 0 otherwise. Let R_d_ = 1 if a person has received a positive test result by date d and R_d_ = 0 otherwise. Observe that T_d_ = 0 ➪ R_d_ = 0 and R_d_ = 1 ➪ T_d_ = 1. By the Law of Total Probability, the infection rate may be written as follows:

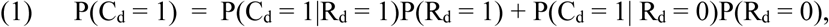

where

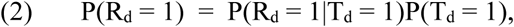

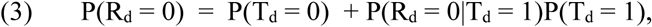

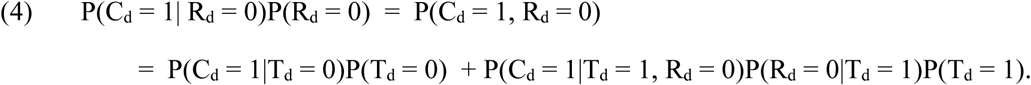

Now consider each of the component quantities that together determine the infection rate. Assuming that reporting of testing is accurate, daily surveillance reveals the testing rate and the rate of positive results among those tested. Thus, the quantities P(T_d_ = 0), P(T_d_ = 1), P(R_d_ = 0|T_d_ = 1), and P(R_d_ = 1|T_d_ = 1) are directly observable. The remaining quantities are not directly observable.

The quantities P(C_d_ = 1|R_d_ = 1) and P(C_d_ = 1|T_d_ = 1, R_d_ = 0) are determined by the accuracy of testing. The former is the positive predictive value (PPV) and the latter is one minus the negative predictive value (NPV). We note that medical researchers and clinicians often measure test accuracy in a different way, through test sensitivity and specificity. The sensitivity and specificity of tests for COVID-19 on the tested sub-population are P(R_d_ = 1|T_d_ = 1, C_d_ = 1) and P(R_d_ = 0|T_d_ = 1, C_d_ = 0) respectively. Sensitivity and specificity are related to PPV and NPV through Bayes Theorem, whose application generally requires knowledge of P(C_d_ = 1|T_d_ = 1), the infection rate in the tested sub-population. An exception to this generalization is that PPV equals one if specificity equals one, whenever sensitivity is positive and P(C_d_ = 1|T_d_ = 1) > 0.^4^

Medical experts believe that the PPV of the prevalent tests for COVID-19 is close to one, but that NPV may be considerably less than one. We have obtained this information in part from personal communication with an infectious disease specialist at Northwestern Memorial Hospital and in part from the public literature. For example, USA Today has reported as follows:^5^

> “Dwayne Breining, executive director for Northwell Labs in New Hyde Park, New York, said the test is extremely accurate and can detect even low levels of the virus. False positives are highly unlikely, he said, though false negatives may result from poor-quality swabs or if the instrument is blocked by mucus. Those factors might have been at play in a number of false negatives initially reported. Patients who continue to have symptoms after a negative test are advised to get retested.”

We therefore find it credible to assume that P(C_d_ = 1|R_d_ = 1) = 1. It can be shown that this is equivalent to assuming that test specificity P(R_d_ = 0|T_d_ = 1, C_d_ = 0) = 1. The final sentence of the Breining quote explains part of why it may not be completely accurate to assume that persons are tested at most once. Another reason is that hospitalized patients are tested to verify recovery before they are released from the hospital. Nevertheless, we maintain this assumption for simplicity.

There does not appear to presently be a firm basis to determine the precise NPV of the prevalent nasal-swab tests, but there may be a basis to determine a credible bound. Medical experts have been cited as believing that the rate of false-negative test findings is at least 0.3. However, it is not clear whether they have in mind one minus the NPV or one minus test sensitivity.^6^ One may perhaps find it credible to extrapolate from experience testing for influenza to testing for covid-19. For example, Peci *et al*. (2014) study the performance of rapid influenza diagnostic testing. They find a PPV of 0.995 and an NPV of 0.853.

It is not clear whether NPV has been constant over the short time period we study or, contrariwise, has varied as testing methods and the subpopulation of tested persons change over time.^7^ The NPV may also vary over longer periods if the virus mutates significantly. The illustrative results that we report later assume that NPV is in the range [0.6, 0.9], implying that P(C_d_ = 1|T_d_ = 1, R_d_ = 0) ∊ [0.1, 0.4].^8^

It remains to consider P(C_d_ = 1|T_d_ = 0), the rate of infection among those who have not been tested. This quantity has been the subject of much discussion, with substantial uncertainty expressed about its value. It may be that the value changes over time as criteria for testing people evolve and testing becomes more common. The illustrative results that we report later show numerically how the conclusions one can draw about P(C_d_ = 1) depend on the available knowledge of P(C_d_ = 1|T_d_ = 0).

To finalize the logical derivation of a bound on P(C_d_ = 1), let [L_d0_, U_d0_] and [L_d10_, U_d10_] denote credible lower and upper bounds on P(C_d_ = 1|T_d_ = 0) and P(C_d_ = 1|T_d_ = 1, R_d_ = 0) respectively. Now combine these bounds with the assumption that P(C_d_ = 1|R_d_ = 1) = 1 and with empirical knowledge of the testing rate and the rate of positive test results. Then equations (1) – (4) imply this bound on the population infection rate:

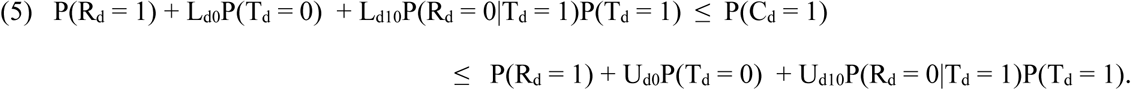

The width of bound (5) is

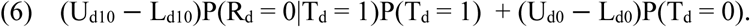

Inspection of (6) shows that uncertainty about test accuracy and about the infection rate in the untested sub-population, measured by U_d10_ − L_d10_ and U_d0_ − L_d0_, combine linearly to yield uncertainty about the population infection rate. The fractions P(T_d_ = 1) and P(T_d_ = 0) of the population who have and have not been tested linearly determine the relative contributions of the two sources of uncertainty.

### 2.2. Using Monotonicity Assumptions to Bound the Infection Rate of Untested Persons

As of April 2020, the fraction of the population who have been tested is very small in most locations. For example, the fraction who have been tested by April 24, 2020 was about 0.015 in Illinois, 0.04 in New York, and 0.027 in Italy; see Section 3 for details on the data sources. Hence, the present dominant concern is uncertainty about the infection rate in untested sub-populations. We now consider the problem of obtaining a credible bound on this quantity.

The worst case from the perspective of uncertainty would occur if society possesses no credible information about the value of P(C_d_ = 1|T_d_ = 0), so U_d0_ − L_d0_ = 1. Then the bound on the population infection rate has width no smaller than P(T_d_ = 0), even if test accuracy is known precisely. In the three locations and on the date mentioned above, the widths of the bounds on the population infection rate are greater than (0.985, 0.960, 0.973). Thus, the available data on the rate of positive tests for tested persons reveal almost nothing about the population infection rate. Moreover, a huge increase in the rate of testing would be required to substantially narrow the width of the bound.

The best feasible case would occur with random testing of a large enough sample of persons to make statistical imprecision a negligible concern. Then P(C_d_ = 1) = P(C_d_ = 1|T_d_ = 1) and uncertainty stems only from incomplete knowledge of the NPV of testing. The Law of Total Probability, the maintained assumption that positive test results are always accurate, and the specified bounds [L_d10_, U_d10_] on P(C_d_ = 1|T_d_ = 1, R_d_ = 0) yield

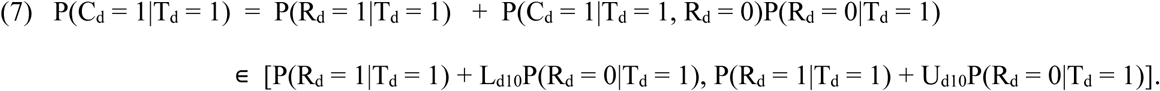

This bound has width (U_d10_ − L_d10_)P(R_d_ = 0|T_d_ = 1).

We judge the current situation to be intermediate between the worst and best case scenarios. We thus far have selective rather than random testing. On the other hand, we have information about the nature of the selectivity, so it is too pessimistic to view society as knowing nothing about infection in the untested sub-population. Two monotonicity assumptions are highly credible in the current context.

#### Testing Monotonicity

Present criteria for testing persons for infection by the coronavirus commonly require the person to display symptoms of infection or to have been in close contact with someone who has tested positive. These criteria strongly suggest that, as of each date d, the infection rate among tested persons is higher than the rate among untested persons. This yields the *testing-monotonicity* assumption

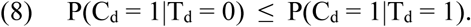

Observe that if testing for infection were random rather than determined by the current criteria, it would be credible to impose a much stronger assumption, namely P(C_d_ = 1|T_d_ = 0) = P(C_d_ = 1|T_d_ = 1). However, testing clearly has not been random. Hence, we only impose assumption (8).

Research on partial identification has often exploited monotonicity assumptions similar to (8), beginning with Manski and Pepper (2000). Combining (8) with the upper bound on P(C_d_ = 1|T_d_ = 1) in (7) yields this upper bound on P(C_d_ = 1|T_d_ = 0).

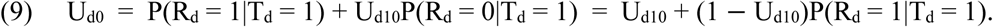

Bound (9) is methodologically interesting because U_d0_ is now a function of U_d10_ rather than a separate quantity. It thus enhances the importance of securing an informative upper bound on P(C_d_ = 1|T_d_ = 1, R_d_ = 0). In particular, (9) implies that U_d0_ ≥ U_d10_, whatever the rate P(R_d_ = 1|T_d_ = 1) of positive test outcomes may be.

The monotonicity assumption does not affect the lower bound L_d0_, which is zero in the absence of other information. Hence, inserting L_d0_ = 0 and (9) into the bound (5) on P(C_d_ = 1) yields

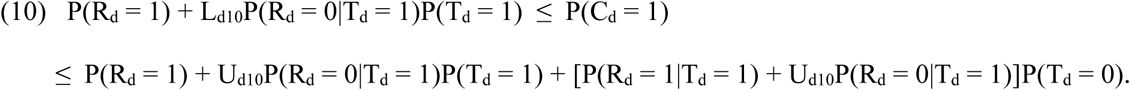

The width of bound (10) is

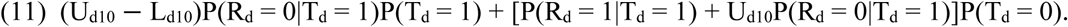

In the present context where P(T_d_ = 1) is very small, the width of the bound approximately equals the sum of the rate P(R_d_ = 1|T_d_ = 1) of positive test results plus the product of the rate P(R_d_ = 0|T_d_ = 1) of negative test results and the upper bound on P(C_d_ = 1|T_d_ = 1, R_d_ = 0).

Bound (10) assumes that a person is tested at most once by date d. If some persons are tested multiple times, publicly reported count data for “total tested” overestimates P(T_d_ = 1). In that case, the bound is too small. To see this, rewrite expression (10) as^9^

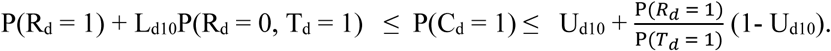

Observe that, if the event T_d_ = 1 sometimes occurs because the same person is tested multiple times, the lower bound is too high and the upper bound is too low.

#### Temporal Monotonicity

A second form of monotonicity holds logically rather than by assumption. Our analysis thus far has only considered the infection rate by a specified date. A person who has been infected by an early date necessarily has been infected by every later date. Hence, for two dates d and d’, we have the *temporal monotonicity condition*

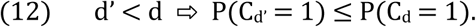

Inequality (12) makes date a monotone *instrumental variable* as defined in Manski and Pepper (2000). Proposition 1 of that article shows that, given a set of date-specific lower and upper bounds on the infection rate for various dates, condition (12) implies that P(C_d_ = 1) must be greater than or equal to the maximum of the date-specific lower bounds for all d’ ≤ d. Moreover, P(C_d_ = 1) must be less than or equal to the minimum of the date-specific upper bounds for all d’ ≥ d.^10^ Applying this result to the date-specific bounds (10) yields this result:

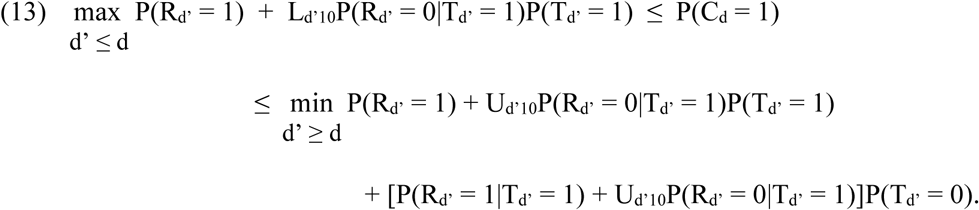

Bound (13) necessarily is a subset of bound (10). The lower bound in (13) equals the lower bound in (10) if the lower bound in (10) at all dates d’ < d is always less than or equal to the lower bound at d. If this does not hold, as can happen with some configurations of testing data, the lower bound on (13) is greater than the one in (10). Symmetrically, the upper bound in (13) equals the upper bound (10) if the upper bound in (10) at all dates d’ > d is always greater than or equal to the upper bound at d. If this does not hold, the upper bound on (13) is less than the one in (10). Thus, the temporal monotonicity condition may or may not have identifying power, depending on the testing data. We find that it modestly improves lower bounds with the data we use.

### 2.3. Bounding the Fraction of Asymptomatic Infections

We are presently unaware of other assumptions or logical conditions that enjoy credibility comparable to the above monotonicity assumptions and that have identifying power. One may, however, perhaps feel comfortable bringing to bear assumptions whose credibility stems from limited evidence or from the judgement of respected medical and epidemiological experts. We provide an example here to illustrate how this may be done and the identifying power studied. We do not endorse the specific assumptions made here.

Consider the decomposition of COVID-19 episodes into those where the patient does and does not manifest discernible symptoms. Dr. Anthony Fauci, the director of the National Institute of Allergy and Infectious Diseases, has been quoted as saying that the fraction of cases in which the patient is infected but shows no symptoms is “somewhere between 25 and 50 percent.” Fauci went on to say “And trust me, that is an estimate. I don’t have any scientific data yet.”^11^

Supposing it to be correct, Fauci’s bound has identifying power when combined with a further assumption. Let A_d_ = 1 or S_d_ = 1 if a person has respectively had an asymptomatic or symptomatic case of COVID-19 by date d. Let each quantity equal zero otherwise. The two categories of illness are mutually exclusive, so C_d_ = A_d_ + S_d_. Hence,

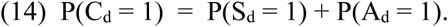

Fauci imposes the assumption

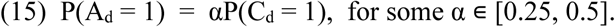

Combining (14) and (15) yields

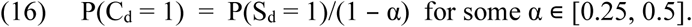

The value of P(S_d_ = 1) is unknown. However, the existing criteria for testing require the presence of symptoms or close contact with a person known to have been infected. Suppose that persons who are accepted for testing all meet the first criterion. Then the rate of infection with symptoms is greater than or equal to the fraction of the population who are both tested and infected. Assume as earlier that all positive testing results are accurate and that the fraction of inaccurate negative testing results is in the bound [L_d10_, U_d10_]. Then the fraction who are tested and infected is known to be at least equal to P(R_d_ = 1) + L_d10_P(T_d_ = 1, R_d_ = 0). Combining this lower bound on P(S_d_ = 1) with knowledge that α ≥ 0.25, (16) yields this lower bound on the population infection rate:

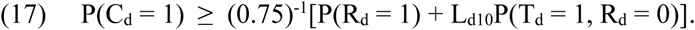

This lower bound is (0.75)^-1^ times that in (10), thus improving on it. If one finds bound (17) credible, the final lower bound on P(C_d_ = 1) is the maximum of the lower bounds in (17) across dates d’ ≤ d, as in (13).

A substantial increase in lower bound (17) results if, instead of relying on Dr. Fauci’s judgment, one brings to bear limited but suggestive evidence on the fraction of asymptomatic infections. Sutton *et al*. (2020) report the findings of universal testing of 215 pregnant women who were admitted for infant delivery to a New York City hospital in late March and early April 2020. The women were screened for symptoms on admissions and were tested. It was found that 29 of the 33 patients who tested positive (87.9%) had no symptoms of Covid-19. If one finds it credible to assume that this hospital-specific and subpopulation-specific^12^ finding holds in general, then α = .879 and the lower bound in (17) increases to

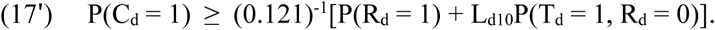

We also find it noteworthy that the rate of confirmed cases in the hospital was 33/215 = 0.153. With universal testing, this finding does not suffer from the huge missing data problem that plagues population statistics on infection. However, it may under-estimate the true infection rate due to test inaccuracy. The implication for inference on the fraction of asymptomatic infections is uncertain.

A lower bound intermediate between (17) and (17’) results if one brings to bear evidence from a recent study of data collected in the small Italian town Vò, where 85.9% of the resident population (for a total of 2,812 individuals) were tested for the presence of SARS-CoV-2 infection in nasal swabs at the early stages of the pandemic (on February 23^rd^), and then 71.5% (for a total of 2,343 individuals) were retested after fourteen days of lockdown. Lavezzo *et al*. (2020) report that 43.2% of the confirmed infections detected across the two surveys were asymptomatic. Using this evidence yields

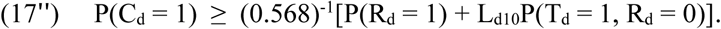

### 2.4. Bounding Rates of Severe Illness Conditional on Infection

Surveillance systems may report several rates of severe illness (V), including hospitalization (H), ICU usage (U), and death (D).^13^ The present discussion considers these reports to be accurate. Thus, one may have empirical knowledge of the rates P(V_d_ = 1) for V ∊ {H, U, D}.

Surveillance systems do not report rates of severe illness conditional on infection. These have the form

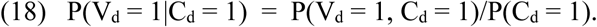

The numerator P(V_d_ = 1, C_d_ = 1) may logically differ from the reported rate P(V_d_ = 1). This may occur for H and U if some persons hospitalized for COVID-19 are mis-diagnosed. It may occur for D if some reported causes of death are inaccurate. For simplicity, we assume here that such errors do not occur. However, we caution that the assumption may not be realistic.^14^

In the absence of reporting errors, V_d_ = 1 ➪ C_d_ = 1. Hence,

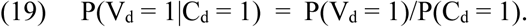

Given (19), the bound obtained for P(C_d_ = 1) immediately yields a bound for P(V_d_ = 1|C_d_ = 1). The lower (upper) bound on P(V_d_ = 1|C_d_ = 1) is achieved when P(C_d_ = 1) takes it upper (lower) bound.

When interpreting rates of severe illness conditional on infection, one should keep in mind that severe cases of COVID-19 may not be apparent as of the date of infection. Many patients begin with mild or no symptoms and develop severe cases a week to two weeks after infection. Hence, the rate of severe illness computed as of a specified date may understate the rate of eventual severe illness.

### 2.5. Bounding Rates Conditional on Personal Characteristics

The above derivations hold however one defines the population of interest. Application of the bound on the infection rate is possible if one has empirical knowledge of the testing rate and the rate of positive testing findings for the relevant population. Application of the bound on the rate of severe illness conditional on infection is possible if one additionally has knowledge of the rate of severe illness in the relevant population.

There are both clinical and public health reasons why one would like to know P(C_d_ = 1|X) and P(V_d_ = 1|X, C_d_ = 1) for persons with specified personal characteristics X. For example, it has been thought important to know these rates conditional on the demographic characteristics X = (age, gender, race). Whatever X may be, the bound on P(C_d_ = 1|X) is this X-specific version of (5):

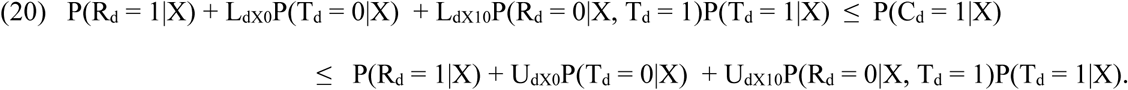

When computing the bound, one should bring to bear credible X-specific bounds on P(C_d_ = 1|X, T_d_ = 0) and P(C_d_ = 1|X, T_d_ = 1, R_d_ = 0). If one imposes a monotonicity restriction conditional on X as in Section 2.2, the bound in (10) is updated similarly as we did in (14) for the bound in (5). The bound on P(V_d_ = 1|X, C d = 1) is computable if surveillance additionally reports P(V_d_ = 1|X).

## 3. Data

We analyze data from two states in the United States, Illinois and New York, and from Italy. Our primary data sources are Illinois Department of Public Health (2020), New York State Department of Health (2020), and the Italian Protezione Civile (2020). The data report cumulative counts of the number of tests performed and the number of persons with positive test results for each day, as well as death counts, starting February 24 for Italy, March 10 for Illinois, and March 2 for New York^15^ ^16^ Our analysis begins from the first day after which all of Illinois, New York, and Italy, had at least one hundred confirmed cases, which is March 16, and runs until April 24. For Illinois and New York, we obtain population size counts using data from Census Bureau (2019) as of July 1, 2019. For Italy, population size is taken from Istituto Nazionale di Statistica (2019) as of January 1, 2019. Our estimates of the quantities in bound (10) are obtained as simple frequency estimators. For example, P(T_d_ = 1) is computed as the number of tests performed up to date d divided by population size.

## 4. Results

Table 1 reports, in columns 2-7, publicly released values of P(T_d_ = 1) and P(R_d_ = 1|T_d_ = 1) for Illinois, New York, and Italy. For several reasons, we caution against comparison of these quantities across states and countries. The criteria for testing and the accuracy of tests may differ across location and may have changed over time. Moreover, the epidemics began at different dates across locations.

**Table 1:** Probability of being tested, of receiving a positive test result if tested, and of death.

| Date | Illinois |  |  | New York |  |  | Italy |  |  |
| --- | --- | --- | --- | --- | --- | --- | --- | --- | --- |
| | $P(T_d=1)$ | $P(R_d=1\vert T_d=1)$ | $P(D_d=1)$ | $P(T_d=1)$ | $P(R_d=1\vert T_d=1)$ | $P(D_d=1)$ | $P(T_d=1)$ | $P(R_d=1\vert T_d=1)$ | $P(D_d=1)$ |
| 3/16/2020 | 0.000 | 0.092 | 0.00000 | 0.001 | 0.134 | 0.00000 | 0.002 | 0.203 | 0.00004 |
| 3/17/2020 | 0.000 | 0.107 | 0.00000 | 0.001 | 0.161 | 0.00000 | 0.002 | 0.212 | 0.00004 |
| 3/18/2020 | 0.000 | 0.140 | 0.00000 | 0.001 | 0.184 | 0.00000 | 0.003 | 0.216 | 0.00005 |
| 3/19/2020 | 0.000 | 0.134 | 0.00000 | 0.002 | 0.218 | 0.00000 | 0.003 | 0.225 | 0.00006 |
| 3/20/2020 | 0.000 | 0.136 | 0.00000 | 0.002 | 0.226 | 0.00000 | 0.003 | 0.227 | 0.00007 |
| 3/21/2020 | 0.000 | 0.121 | 0.00000 | 0.003 | 0.246 | 0.00000 | 0.004 | 0.230 | 0.00008 |
| 3/22/2020 | 0.001 | 0.125 | 0.00000 | 0.004 | 0.266 | 0.00001 | 0.004 | 0.229 | 0.00009 |
| 3/23/2020 | 0.001 | 0.130 | 0.00000 | 0.005 | 0.280 | 0.00001 | 0.005 | 0.232 | 0.00010 |
| 3/24/2020 | 0.001 | 0.134 | 0.00000 | 0.005 | 0.297 | 0.00001 | 0.005 | 0.233 | 0.00011 |
| 3/25/2020 | 0.001 | 0.131 | 0.00000 | 0.006 | 0.305 | 0.00001 | 0.005 | 0.229 | 0.00012 |
| 3/26/2020 | 0.001 | 0.153 | 0.00000 | 0.007 | 0.322 | 0.00002 | 0.006 | 0.223 | 0.00014 |
| 3/27/2020 | 0.002 | 0.140 | 0.00000 | 0.008 | 0.335 | 0.00003 | 0.007 | 0.219 | 0.00015 |
| 3/28/2020 | 0.002 | 0.137 | 0.00000 | 0.009 | 0.345 | 0.00004 | 0.007 | 0.215 | 0.00017 |
| 3/29/2020 | 0.002 | 0.166 | 0.00001 | 0.010 | 0.356 | 0.00005 | 0.008 | 0.215 | 0.00018 |
| 3/30/2020 | 0.002 | 0.166 | 0.00001 | 0.011 | 0.369 | 0.00006 | 0.008 | 0.213 | 0.00019 |
| 3/31/2020 | 0.003 | 0.170 | 0.00001 | 0.011 | 0.379 | 0.00008 | 0.008 | 0.209 | 0.00021 |
| 4/1/2020 | 0.003 | 0.173 | 0.00001 | 0.012 | 0.387 | 0.00010 | 0.009 | 0.204 | 0.00022 |
| 4/2/2020 | 0.003 | 0.176 | 0.00001 | 0.013 | 0.395 | 0.00012 | 0.010 | 0.198 | 0.00023 |
| 4/3/2020 | 0.004 | 0.185 | 0.00002 | 0.015 | 0.401 | 0.00015 | 0.010 | 0.193 | 0.00024 |
| 4/4/2020 | 0.004 | 0.193 | 0.00002 | 0.016 | 0.404 | 0.00018 | 0.011 | 0.190 | 0.00025 |
| 4/5/2020 | 0.005 | 0.191 | 0.00002 | 0.016 | 0.407 | 0.00021 | 0.011 | 0.186 | 0.00026 |
| 4/6/2020 | 0.005 | 0.195 | 0.00002 | 0.017 | 0.408 | 0.00024 | 0.012 | 0.184 | 0.00027 |
| 4/7/2020 | 0.005 | 0.197 | 0.00003 | 0.019 | 0.409 | 0.00028 | 0.013 | 0.179 | 0.00028 |
| 4/8/2020 | 0.006 | 0.201 | 0.00004 | 0.020 | 0.408 | 0.00032 | 0.013 | 0.173 | 0.00029 |
| 4/9/2020 | 0.006 | 0.203 | 0.00004 | 0.021 | 0.408 | 0.00036 | 0.014 | 0.168 | 0.00030 |
| 4/10/2020 | 0.007 | 0.204 | 0.00005 | 0.023 | 0.409 | 0.00040 | 0.015 | 0.163 | 0.00031 |
| 4/11/2020 | 0.007 | 0.207 | 0.00005 | 0.024 | 0.409 | 0.00044 | 0.016 | 0.158 | 0.00032 |
| 4/12/2020 | 0.008 | 0.207 | 0.00006 | 0.025 | 0.408 | 0.00048 | 0.017 | 0.155 | 0.00033 |
| 4/13/2020 | 0.008 | 0.208 | 0.00006 | 0.026 | 0.405 | 0.00052 | 0.017 | 0.152 | 0.00034 |
| 4/14/2020 | 0.009 | 0.210 | 0.00007 | 0.027 | 0.406 | 0.00056 | 0.018 | 0.151 | 0.00035 |
| 4/15/2020 | 0.009 | 0.210 | 0.00007 | 0.028 | 0.404 | 0.00060 | 0.019 | 0.148 | 0.00036 |
| 4/16/2020 | 0.010 | 0.210 | 0.00008 | 0.029 | 0.401 | 0.00063 | 0.020 | 0.143 | 0.00037 |
| 4/17/2020 | 0.010 | 0.212 | 0.00009 | 0.031 | 0.397 | 0.00066 | 0.021 | 0.139 | 0.00038 |
| 4/18/2020 | 0.011 | 0.212 | 0.00010 | 0.032 | 0.393 | 0.00069 | 0.022 | 0.135 | 0.00038 |
| 4/19/2020 | 0.011 | 0.212 | 0.00010 | 0.033 | 0.390 | 0.00071 | 0.022 | 0.132 | 0.00039 |
| 4/20/2020 | 0.012 | 0.212 | 0.00011 | 0.033 | 0.388 | 0.00074 | 0.023 | 0.130 | 0.00040 |
| 4/21/2020 | 0.012 | 0.213 | 0.00012 | 0.034 | 0.384 | 0.00076 | 0.024 | 0.127 | 0.00041 |
| 4/22/2020 | 0.013 | 0.214 | 0.00012 | 0.036 | 0.379 | 0.00079 | 0.025 | 0.124 | 0.00042 |
| 4/23/2020 | 0.014 | 0.213 | 0.00013 | 0.038 | 0.372 | 0.00081 | 0.026 | 0.120 | 0.00042 |
| 4/24/2020 | 0.015 | 0.209 | 0.00014 | 0.040 | 0.363 | 0.00083 | 0.027 | 0.118 | 0.00043 |

Notes: Population sizes are 12,671,821 for Illinois, 19,453,561 for New York, and 60,359,546 for Italy.

The table reveals that from March 16 to April 24, 2020, the fraction of individuals tested increased from 0 to 0.015 in Illinois, from 0.001 to 0.04 in New York, and from 0.002 to 0.027 in Italy. Over the same period, the fraction of positive test results varied (non-monotonically) from 0.092 to 0.209, with a peak of 0.214, in Illinois; from 0.134 to 0.363, with a peak of 0.409, in New York; and from 0.203 to 0.118 in Italy, with a peak of 0.233. The fraction of individuals who die increased from 0 to 0.00014 for Illinois, from 0 to 0.00083 for New York, and from 0.0004 to 0.00043 for Italy.

Table 2 reports the bounds in (13) for Illinois, New York, and Italy, under the monotonicity assumptions presented in Section 2.2. The temporal monotonicity condition is reflected in the fact that, for each state and country, both the lower and upper bounds on P(C_d_ = 1) weakly increase over time. The widths of the bounds varied (non-monotonically) from 0.455 to 0.521 for Illinois, with a peak of 0.523; 0.48 to 0.601 for New York, with a peak of 0.613; and remained about 0.47 for Italy throughout the period.

**Table 2:** Bounds on infection rate under the testing and temporal monotonicity assumptions.

| Date | Illinois |  | New York |  | Italy |  |
| --- | --- | --- | --- | --- | --- | --- |
|  | LB | UB | LB | UB | LB | UB |
| 3/16/2020 | 0.000 | 0.455 | 0.000 | 0.480 | 0.001 | 0.471 |
| 3/17/2020 | 0.000 | 0.464 | 0.000 | 0.497 | 0.001 | 0.471 |
| 3/18/2020 | 0.000 | 0.472 | 0.000 | 0.511 | 0.001 | 0.471 |
| 3/19/2020 | 0.000 | 0.472 | 0.000 | 0.531 | 0.001 | 0.471 |
| 3/20/2020 | 0.000 | 0.472 | 0.001 | 0.536 | 0.001 | 0.471 |
| 3/21/2020 | 0.000 | 0.472 | 0.001 | 0.547 | 0.001 | 0.471 |
| 3/22/2020 | 0.000 | 0.475 | 0.001 | 0.559 | 0.001 | 0.471 |
| 3/23/2020 | 0.000 | 0.478 | 0.002 | 0.568 | 0.001 | 0.471 |
| 3/24/2020 | 0.000 | 0.479 | 0.002 | 0.578 | 0.002 | 0.471 |
| 3/25/2020 | 0.000 | 0.479 | 0.002 | 0.583 | 0.002 | 0.471 |
| 3/26/2020 | 0.000 | 0.482 | 0.003 | 0.593 | 0.002 | 0.471 |
| 3/27/2020 | 0.000 | 0.482 | 0.003 | 0.601 | 0.002 | 0.471 |
| 3/28/2020 | 0.000 | 0.482 | 0.004 | 0.607 | 0.002 | 0.471 |
| 3/29/2020 | 0.001 | 0.499 | 0.004 | 0.614 | 0.002 | 0.471 |
| 3/30/2020 | 0.001 | 0.500 | 0.005 | 0.618 | 0.002 | 0.471 |
| 3/31/2020 | 0.001 | 0.502 | 0.005 | 0.618 | 0.002 | 0.471 |
| 4/1/2020 | 0.001 | 0.504 | 0.006 | 0.618 | 0.003 | 0.471 |
| 4/2/2020 | 0.001 | 0.506 | 0.006 | 0.618 | 0.003 | 0.471 |
| 4/3/2020 | 0.001 | 0.511 | 0.007 | 0.618 | 0.003 | 0.471 |
| 4/4/2020 | 0.001 | 0.515 | 0.007 | 0.618 | 0.003 | 0.471 |
| 4/5/2020 | 0.001 | 0.515 | 0.008 | 0.618 | 0.003 | 0.471 |
| 4/6/2020 | 0.001 | 0.517 | 0.008 | 0.618 | 0.003 | 0.471 |
| 4/7/2020 | 0.002 | 0.518 | 0.009 | 0.618 | 0.003 | 0.471 |
| 4/8/2020 | 0.002 | 0.521 | 0.009 | 0.618 | 0.003 | 0.471 |
| 4/9/2020 | 0.002 | 0.522 | 0.010 | 0.618 | 0.004 | 0.471 |
| 4/10/2020 | 0.002 | 0.523 | 0.011 | 0.618 | 0.004 | 0.471 |
| 4/11/2020 | 0.002 | 0.524 | 0.011 | 0.618 | 0.004 | 0.471 |
| 4/12/2020 | 0.002 | 0.524 | 0.011 | 0.618 | 0.004 | 0.471 |
| 4/13/2020 | 0.002 | 0.525 | 0.012 | 0.618 | 0.004 | 0.471 |
| 4/14/2020 | 0.003 | 0.525 | 0.013 | 0.618 | 0.004 | 0.471 |
| 4/15/2020 | 0.003 | 0.525 | 0.013 | 0.618 | 0.004 | 0.471 |
| 4/16/2020 | 0.003 | 0.525 | 0.014 | 0.618 | 0.004 | 0.471 |
| 4/17/2020 | 0.003 | 0.525 | 0.014 | 0.618 | 0.005 | 0.471 |
| 4/18/2020 | 0.003 | 0.525 | 0.014 | 0.618 | 0.005 | 0.471 |
| 4/19/2020 | 0.003 | 0.525 | 0.015 | 0.618 | 0.005 | 0.471 |
| 4/20/2020 | 0.003 | 0.525 | 0.015 | 0.618 | 0.005 | 0.471 |
| 4/21/2020 | 0.004 | 0.525 | 0.015 | 0.618 | 0.005 | 0.471 |
| 4/22/2020 | 0.004 | 0.525 | 0.016 | 0.618 | 0.005 | 0.471 |
| 4/23/2020 | 0.004 | 0.525 | 0.016 | 0.618 | 0.005 | 0.471 |
| 4/24/2020 | 0.004 | 0.525 | 0.017 | 0.618 | 0.006 | 0.471 |

The substantial width of the bounds reflects the fact, previously discussed, that the fraction of tested individuals was very small throughout the period. Nonetheless, the bounds have substantial informational content relative to the extremely wide bounds that would hold if society were to possess no credible information about the infection rate among untested persons. On April 24, 2020, the bounds on the infection rates in Illinois, New York, and Italy respectively are [0.004, 0.525], [0.017, 0.618], and [0.006, 0.471].

We next bring to bear information on the rate of asymptomatic infections as discussed in Section 2.3. This information does not lower the upper bound on the probability of infection, but it raises the lower bounds. Using the expert opinion of Dr. Fauci, the lower bounds increase by a factor of (0.75)^-1^. Considering again April 24, 2020, the updated bounds are [0.006, 0.525], [0.023, 0.618], and [0.007, 0.471]. Using the evidence from the city of Vò in Italy, the lower bounds increase by (0.568)^-1^. These updated bounds are [0.008, 0.525], [0.03, 0.618], and [0.01, 0.471]. Using the evidence from the hospital in New York City, the lower bounds increase by (0.121)^-1^. These updated bounds are [0.036, 0.525], [0.141, 0.618], and [0.046, 0.471].

Table 3 reports the probability that infected individuals die (the case fatality ratio), under the monotonicity assumptions for infection rates presented in Section 2.2. Focusing on April 24, 2020, we see that the bounds are [0, 0.033] for Illinois, [0.001, 0.049] for New York, and [0.001, 0.077] for Italy. It is notable that the upper bounds on fatality are substantially lower than the fatality rate among confirmed infected individuals, which were, respectively, 0.045, 0.059, and 0.134 on April 24.

**Table 3:**
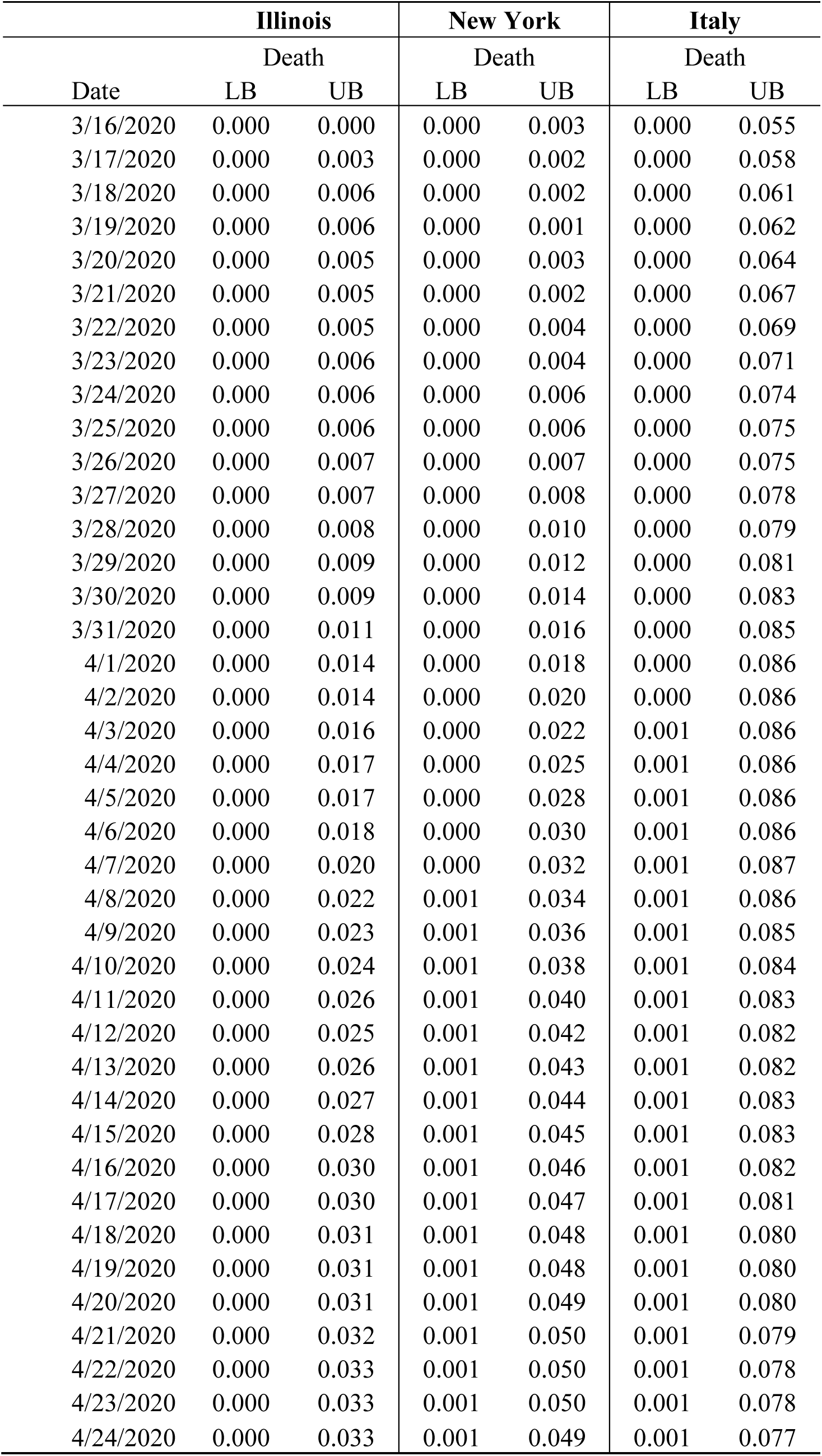
Bounds on the probability of death conditional on infection under the testing and temporal monotonicity assumptions.

If one brings to bear information on the rate of asymptomatic infections as discussed in Section 2.3, these upper bounds further shrink substantially. Using the expert opinion of Dr. Fauci, the upper bounds are multiplied by a factor of 0.75. Considering again April 24, 2020, the updated bounds are [0, 0.025], [0.001, 0.037], and [0.001, 0.058]. Using the evidence from the city of Vò in Italy, the upper bounds are multiplied by 0.568. The updated bounds are [0, 0.019], [0.001, 0.028], and [0.001, 0.044]. Using the evidence from the hospital in New York City, the upper bounds are multiplied by 0.121. These updated bounds are [0, 0.004], [0.001, 0.006], and [0.001, 0.009].

## 5. Discussion

This paper has used standard methods of partial identification analysis to study two key aspects of the uncertainty that has frustrated attempts to learn the COVID-19 cumulative infection rate and rates of severe illness conditional on infection. We have quantified the implications of uncertainty about the infection rate among non-tested persons and about the NPV of the tests in use. The simple analysis of Section 2 shows how available data and maintained assumptions combine to determine the inferences that can logically be drawn. We have used monotonicity assumptions that have strong credibility in the current context. We also have used suggestive information on the rate of asymptomatic infection to illustrate how further assumptions having a less firm foundation may be brought to bear, should one find them credible.

We have used data for two American states and for Italy to illustrate application of the analysis. Given that the tested fraction of the population has been very low, one can barely draw any conclusion about the population infection rate without making assumptions that bound the rate of infection in the untested sub-population. Imposing the monotonicity assumptions restricts the population infection rate to bounds that have about width 0.5 in the current COVID-19 context.

One naturally may prefer bounds of narrower width. Given the available data, this is logically possible to achieve only if one imposes stronger assumptions with considerable identifying power. We have not reported narrower bounds because we do not immediately see a credible basis to add assumptions that would justify them. Readers who feel that they can motivate stronger assumptions may adapt our analysis to determine their implications.

Among the possibilities for narrowing the bounds that we plan to investigate, it has often been suggested that we can learn about the prevalence and severity of COVID-19 in one location by observing the experiences of populations in other locations. For example, it has been suggested that the United States can learn from the experience in China, South Korea, and Italy. In these locations the epidemic began earlier and has been handled in different ways. Bringing to bear data from different locations is not helpful per se. It may be helpful if the data are combined with assumptions that enable credible extrapolation across locations. Given such assumptions, the partial-identification sub-literature on *intersection bounds* shows how to proceed formally to tighten inference. See Manski (2020) and Molinari (2020).

We also plan to explore imposition of assumptions on the dynamics of the epidemic that have been used in epidemiological modeling and that may have some credibility. For example, a shape restriction commonly maintained in epidemiological models is that the function describing the time-series variation in the rate of new infection is single peaked. Equivalently, this assumption holds that the cumulative rate of infection is S-shaped. This and other shape restrictions may have identifying power.

To simplify the presentation, we have intentionally abstracted from other potential sources of uncertainty that may further aggravate the inferential problem. We have assumed that persons who recover from the COVID-19 disease become immune and, hence, cannot be infected anew. We have assumed that persons are tested at most once. We have assumed that hospitals correctly diagnose patients and that public records correctly code causes of death. We caution that these assumptions may not be completely accurate, as discussed earlier in the paper. The partial identification analysis performed in Section 2 may be extended to incorporate these and other further uncertainties.

Departing from conventional practice in applied econometric analysis, we do not refer to the empirical results in Section 4 as “estimates” and we do not provide measures of statistical precision. Instead, we view states and nations as the units of interest rather than as realizations from some sampling process. Measurement of statistical precision requires specification of a sampling process that generates the available data. Yet we are unsure what type of sampling process would be reasonable to assume in this work.

The data we used are exact population counts of tests performed and their results in each location, not observations of samples drawn in the locations. To perform statistical inference, one would have to view the population of each location as the sampling realization of a random process defined on a super-population of alternative population sizes and compositions. See Manski and Pepper (2018) for extended discussion of this matter in a different applied context.

Topics for future research include inference on the fraction of the population who are currently ill and the daily rate of new infection. A difficulty in studying the former is that the duration of the disease is apparently quite heterogeneous and not well understood. The latter can in principle be bounded once one bounds the cumulative infection rate, as the rate of new infection is the derivative of the cumulative infection rate. It appears that obtaining informative bounds on this derivative requires stronger assumptions than those we have considered.

## 6. Conclusion

While the bounds we report can be narrowed by imposing stronger assumptions, a more satisfactory way to increase knowledge of the infection rate is to obtain better data. As has been widely recognized, random testing of populations would contribute enormously. Random samples need not be huge to be informative, but they should be large enough to make statistical imprecision no more than a modest concern when interpreting findings. Obtaining a firm understanding of the negative predictive value of the tests in use is also important. We urge efforts to progress in both directions.

## Data Availability

The data are publicly available. An Excel file is provided by a link on the author's personal webpage.

http://faculty.wcas.northwestern.edu/~cfm754/covid-19_data.xlsx

## Acknowledgements

We thank Yizhou Kuang for able research assistance. We thank Mogens Fosgerau, Michael Gmeiner, Valentyn Litvin, John Pepper, Jörg Stoye, Elie Tamer, and an anonymous reviewer for helpful comments. We are grateful for the opportunity to present this work at an April 13, 2020 virtual seminar at the Institute for Policy Research, Northwestern University.

## Footnotes

1 https://www.who.int/dg/speeches/detail/who-director-general-s-opening-remarks-at-the-media-briefing-on-covid-19---3-march-2020

2 See, for example, https://www.statnews.com/2020/03/17/a-fiasco-in-the-making-as-the-coronavirus-pandemic-takes-hold-we-are-making-decisions-without-reliable-data/

3 See https://www.imperial.ac.uk/mrc-global-infectious-disease-analysis/covid-19/ and http://www.healthdata.org/ respectively.

4 Given that testing is necessary for a positive test result, Bayes Theorem relates the PPV to sensitivity and specificity as follows: 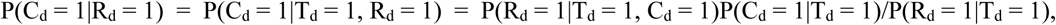 where 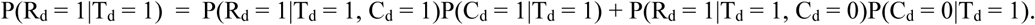 Having specificity equal one is equivalent to P(R_d_ = 1|T_d_ = 1, C_d_ = 0) = 0. Hence, P(C_d_ = 1|R_d_ = 1) = 1 when P(C_d_ = 1|T_d_ = 1) > 0 and P(R_d_ = 1|T_d_ = 1, C_d_ = 1) > 0.

5 https://www.usatoday.com/story/news/nation/2020/03/16/coronavirus-what-expect-when-you-get-tested-covid-19/5061120002/

6 See, for example, https://www.livescience.com/covid19-coronavirus-tests-false-negatives.html.

7 Failure of a test to detect that a person has been infected may occur for multiple reasons. In the current context, where eligibility for testing requires a person to exhibit symptoms or to have been in recent close contact with a confirmed case, we expect that imperfect administration of tests is the dominant reason for inaccuracy in results. In settings with widespread eligibility for testing, inaccuracy also occurs if a person is tested after recovery from COVID-19. Then a nasal swab test would show no presence of the virus and yield a negative result, missing the fact that the person was infected previously.

8 Rather than assume a bound on NPV directly, a medical researcher or clinician could assume a bound on sensitivity. It can be shown that a bound on sensitivity combined with the assumption that specificity equals one implies a bound on the NPV.

9 To obtain the upper bound, use the Law of Total Probability to write 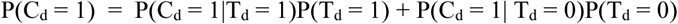 Then combine the upper bound in expression (7) with the one in expression (9).

10 Proposition 1 of Manski and Pepper (2000) shows that this bound is sharp. That is, it is the tightest bound achievable with the available information. Molinari (2020, Section 2.1) shows that it is a more complex matter to obtain sharp bounds for functions of the infection rate that vary with time.

11 See https://www.nytimes.com/2020/04/07/science/coronavirus-uncertainty-scientific-trust.html?action=click&module=Well&pgtype=Homepage&section=Health

12 In addition to this subpopulation being female, pregnancy typically occurs during a limited age range, and we have no information on whether age and gender systematically affect the presence of symptoms conditional on infection.

13 Hospitalization includes ICU usage.

14 See https://www.washingtonpost.com/investigations/coronavirus-death-toll-americans-are-almost-certainly-dying-of-covid-19-but-being-left-out-of-the-official-count/2020/04/05/71d67982-747e-11ea-87da-77a8136c1a6d_story.html

15 In all locations, a person is classified as having a confirmed case of the disease by date d if the person has obtained a positive test result by that date. The New York documentation of testing indicates that non-positive results are sub-classified into those that are negative and inconclusive. This sub-classification is not made in the documentation for Illinois and Italy. Receipt of test results may take several days or longer. We are not certain how agencies classify persons while results are still pending. Our analysis interprets the reported data on confirmed cases to exclude cases where test results are pending. Whereas statistics on cumulative confirmed cases count the number of persons with positive test results to date, statistics on cumulative testing count the number of tests performed. The New York documentation states this: > “Test counts reflect those completed on an individual each day. A person may have multiple specimens tested on one day, these would be counted one time, i.e., if two specimens are collected from an individual at the same time and then evaluated, the outcome of the evaluation of those two samples to diagnose the individual is counted as a single test of one person, even though the specimens may be tested separately. Conversely, if an individual is tested on more than one day, the data will show two tests of an individual, one for each date the person was tested.” Thus, to the extent that persons are retested on different days, the cumulative test counts overstate the cumulative number of persons who have been tested.

16 Our data source for deaths in New York is COVID Tracking Project (2020)

